# Extracellular vesicles as biomarkers for psoriatic arthritis: a systematic review & meta-analysis

**DOI:** 10.64898/2026.06.23.26356353

**Authors:** Timothy Zhang, Firas-Shah Zoha, Christopher Zhu, Jordan Ackerfield, Sophia Wang, Jacklyn Luu, Sarah Ning, Erin Suh, Robert H. Brophy, Derrick M. Knapik, Hash Brown Taha

**Author notes:** **Corresponding author Hash Brown Taha**. **Timothy Zhang:**, **Firas-Shah Zoha:**, **Chris Zhu:**, **Jordan Ackerfield:**, **Jacklyn Luu:**, **Sophia Wang:**, **Sarah Ning:**, **Erin Suh:**, **Robert H. Brophy:**, **Derrick M. Knapik:**, **Hash Brown Taha:**.

## Abstract

**Background:** Psoriatic arthritis (PsA) is an inflammatory condition involving joints, tendon-bone entheses and synovium that can develop in individuals with psoriasis. Early, accurate clinical diagnosis remains difficult. Extracellular vesicles (EVs) carry proteins and miRNAs that

**Methods:** PubMed and Embase were searched from inception through May 21st, 2026, and human studies examining EV-associated protein or miRNA biomarkers in PsA and related psoriatic or inflammatory diseases were included, with risk of bias assessed using a modified Newcastle-Ottawa Scale and diagnostic accuracy summarized using HSROC/BRMA models when data were sufficient.

**Results:** Seven studies met the inclusion criteria, including 119 individuals with PsA (weighted mean age: 49.8 years; 43.7% female), 205 individuals with non-PsA psoriasis (weighted mean age: 46.4 years; female %: NA), 55 controls (weighted mean age: 44.5 years; 38.2% female), and 50 individuals with other inflammatory joint disorders (weighted mean age: 58.0 years; 58.0% female). EV-associated protein markers demonstrated heterogeneous findings related to immune, vascular, inflammatory, and osteoimmunological signaling. Only 4.2% (4/95) of miRNAs were consistently identified across studies comparing PsA with non-PsA psoriasis, with lower overlap (1.5%, 1/67) in studies comparing PsA with controls. ROC meta-analysis suggested preliminary diagnostic potential, particularly for distinguishing PsA from non-PsA psoriasis, although evidence was constrained by small study numbers.

**Conclusions:** EV-associated proteins and miRNAs are potential biomarker candidates for PsA, reflecting inflammatory, vascular, and osteoimmunological processes underlying disease pathophysiology. However, current evidence remains preliminary and limited by small cohorts, methodological heterogeneity, and inconsistent reporting across studies.

## Introduction

Psoriatic arthritis (PsA) is a chronic, immune-mediated disease characterized by joint inflammation with involvement of the synovium and tendon-bone entheses. PsA frequently develops in individuals with psoriasis, although the transition from cutaneous disease to musculoskeletal manifestation remains unpredictable and poorly understood [1]. Delayed diagnosis is common due to heterogeneous clinical presentation and overlap with other inflammatory arthropathies, while being associated with worse radiographic and functional outcomes [1].

Current diagnostic approaches for PsA rely primarily on clinical evaluation and classification criteria, such as the Classification Criteria for Psoriatic Arthritis [2]. However, while CASPAR has high sensitivity and specificity in established disease [2], it is currently limited for early disease detection [1]. Conventional inflammatory markers such as C-reactive protein and erythrocyte sedimentation rate, along with pathognomonic radiographic findings such as pencil-in-cup deformity involving the distal interphalangeal joint and paravertebral ossifications, are non-specific and may be normal in a subset of individuals [1]. These limitations have driven increasing interest in circulating molecular biomarkers that may better reflect underlying disease mechanisms and help detect PsA at an earlier disease stage.

Extracellular vesicles (EVs) are a heterogeneous population of small, membrane-bound vesicles including exosomes and ectosomes [3]. Encapsulated by a phospholipid bilayer, EVs carry proteins, lipids, carbohydrates, and nucleic acids, functioning in intercellular communication as well as cellular waste disposal [4]. In inflammatory joint diseases, EVs have been implicated in processes such as cytokine signaling, immune cell activation and tissue remodeling, including pathways involving tumor necrosis factor-alpha (TNF-α), interluken-6 (IL-6), and nuclear factor-kappa beta (NF-κB) [5–7]. Importantly, EV-associated cargo reflects cell-state-specific messages while being protected from degradation in circulation, making EVs attractive candidates for minimally invasive biomarker discovery [7–9]. In the context of PsA, both EV-associated proteins and miRNAs have been proposed as potential biomarkers capable of detecting disease activity and inflammatory status [9, 10]. Overall, several studies have reported differential EV-associated protein expression and miRNA expression in PsA, highlighting their diagnostic potential. However, inconsistencies such as differences in EV isolation methods, biofluid usage, cohort characteristics across studies remain a major limitation [11–17].

The aim of this study is to systematically review and synthesize current evidence on EV-associated molecular biomarkers in PsA. Our findings suggest that EV-associated proteins and miRNAs may serve as biologically informative biomarkers of inflammatory, vascular, synovial, and osteoimmunological processes in PsA, with potential relevance for distinguishing PsA from RA and non-arthritic psoriasis; however, small cohorts, methodological heterogeneity, limited reproducibility, and insufficient longitudinal evidence currently limit their clinical applicability. (**Figure 1**).

**Figure 1.**
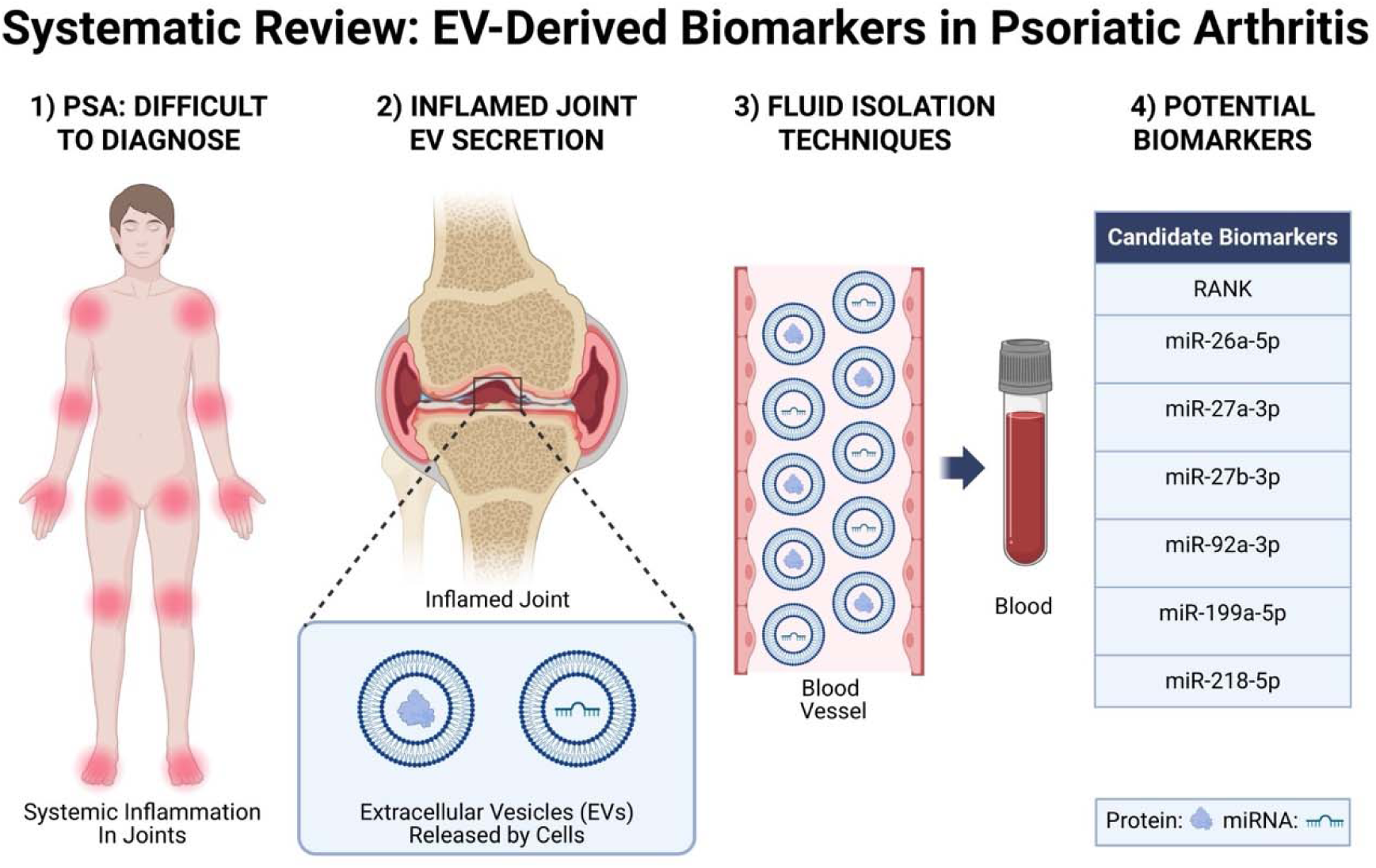
Conceptual overview of extracellular vesicle (EV)-derived biomarkers in psoriatic arthritis (PsA). PsA is a systemic inflammatory disease with musculoskeletal involvement, often presenting diagnostic challenges due to heterogeneous clinical manifestations and overlap with other inflammatory arthropathies. Inflammation within affected joints promotes the release of extracellular vesicles (EVs), membrane-bound particles containing proteins, microRNAs (miRNAs), and other molecular cargo reflective of underlying disease processes. EVs released from inflamed tissues may enter circulation and be isolated from biofluids, including blood, using various EV separation techniques for downstream molecular analysis. Candidate EV-associated biomarkers identified across studies include several dysregulated miRNAs, such as miR-26a-5p, miR-27a-3p, miR-27b-3p, miR-92a-3p, miR-199a-5p, and miR-218-5p.

## Methods

This systematic review was conducted following the guidelines of the Preferred Reporting Items for Systematic Reviews and Meta-Analyses (PRISMA). The study used only anonymized, previously published data and did not involve the collection of personal information or any direct research on human participants; therefore, ethical approval was not required. The study protocol was registered on PROSPERO (CRD420261407481).

### Data sources and search strategy

A thorough search was performed for relevant articles using specific search terms related to PsA and related disorders. The search was conducted in PubMed and EMBASE and covered articles published from the inception of the databases until May 21st, 2026. Two independent researchers (TZ, CZ) screened all titles, abstracts, and full manuscripts to identify articles meeting inclusion criteria. Reference lists of included articles were manually reviewed to identify additional relevant articles, and supplementary searches were conducted using Google Scholar to collect studies not indexed in the primary databases. Any discrepancies in study selection were resolved through consensus discussion with the senior author [initials blinded for peer review], of which no additional studies were identified. The full search strategy can be accessed in (**Table S1)**.

### Eligibility criteria

A comprehensive and intentionally broad search strategy centered on EVs and all subtypes of psoriasis was performed to capture the full scope of relevant articles. Studies were eligible for inclusion if they evaluated EVs in conjunction with PsA, whether psoriatic arthritis patient samples were assessed independently or as a subset of broader portion of individuals with psoriasis.

### Data extraction

Data from all eligible studies were extracted independently by multiple investigators (TZ, FZ, CZ, JA, JL, SN & ES), with cross verification performed to ensure accuracy and completeness. An additional investigator (HBT) reviewed the extracted data for accuracy. Extracted variables included study identifiers (author, year, country, PMID), species, biomarker classification and intended clinical application (diagnostic, prognostic, or predictive), evidence level, preanalytical variables (including fasting status, collection tube type, platelet depletion, lysis/extraction methods, and EV storage conditions), sample source, EV isolation methodologies, EV characterization and confirmation methods, and EV quantification platforms. Cohort level variables included sample sizes for PsA, controls, and other disease groups, demographic variables (age and sex), and disease severity and staging metrics including Psoriasis Area Severity Index (PASI), Disease Activity Score [18], Disease Activity Score 28 (DAS28), Bath Ankylosing Spondylitis Disease Activity Index (BASDAI), body surface area involvement, disease duration, and PsA stage were recorded, where available. Additionally, we extracted whether studies reported quantitative biomarker summary statistics or individual level data visualization, as well as the presence of receiver operating characteristic analyses, correlational analyses, prognostic modeling, predictive modeling, and differential expression analyses for EV associated proteins, cytokines, surface markers, and miRNAs.

For diagnostic analyses, reported AUC, sensitivity, specificity, and sample sizes were extracted when available. When tabulated values were unavailable, WebPlotDigitizer (version 5.2)[19] was used to reconstruct approximate individual-level biomarker values from published scatter plots or ROC coordinates from published ROC curves. Reconstructed biomarker values were analyzed in RStudio (version 4.6.0) using the pROC package; binomial logistic regression was used to model disease status, ROC curves were generated from predicted probabilities, and optimal cutoffs with corresponding sensitivity and specificity were identified using Youden’s index. For studies that provided ROC curves but did not report sensitivity or specificity values, ROC coordinates were digitized using WebPlotDigitizer, and the coordinate maximizing Youden’s index was used to estimate sensitivity and specificity. All digitized values were treated as approximations and documented in supplemental Figures S1-S8. We performed several internal validation procedures for the WebPlotDigitizer-derived data, including extraction from published ROC curves and reconstruction from individualized values when available. In our hands, these approaches yielded near-identical estimates for mean ± SD, AUC, sensitivity, and specificity when compared with the values reported in the original studies, supporting the accuracy and reliability of the digitization process.

### Risk of bias assessment

Risk of bias was assessed using two different modified versions of the Newcastle-Ottawa Scale, one for cross-sectional studies and one for longitudinal studies [20]. Studies were categorized according to the context in which EV biomarkers were evaluated. Specifically, classification was based on whether biomarker measurements occurred cross-sectionally, longitudinally or within a prognostic or predictive framework. Cross-sectional studies that used EV biomarkers as a comparison factor between individuals with PsA and controls or individuals diagnosed with other psoriasis subtypes were grouped as diagnostic studies. Longitudinal studies using EV biomarkers in relation to changes over time, including treatment-response or disease-related associations, were grouped within predictive or prognostic studies. As such, assignment to cross-sectional or longitudinal categories reflected the timing and intended use of EV biomarkers as opposed to the primary design or objectives of the parent study. Risk of bias assessment was conducted by TZ, CZ and JA and further checked by HBT. There were no discrepancies among the scores.

### Data synthesis and statistics

Pooled diagnostic performance was estimated using the hierarchical summary receiver operating characteristic (HSROC) model or the bivariate random-effects meta-analysis (BRMA) model, which are mathematically equivalent when no covariates are included [21]. Models were implemented in Stata using the *metadta* [22] command, and HSROC curves were generated to visualize overall diagnostic performance. Between-study heterogeneity was assessed using the Zhou and Dendukuri I² statistic [23].

## Results

A total of 7 studies [11–17] were identified as meeting inclusion criteria and included in the systematic review (**Figure 2**) of which 4 (27144162, 31954077, 35409365, 39307927) were included in the meta-analysis. All 7 studies (**Table 1**) evaluated EV biomarkers in a cross-sectional and diagnostic context, while one study also examined their predictive value in response to anti-inflammatory treatment [11].The studies included a total of 119 individuals with PsA (weighted mean age: 49.8 years; 52 females; weighted disease duration: 9.2 years; weighted PASI: 8.8; weighted DAS: 3.2), 55 controls (weighted mean age: 44.5 years; 21 females), and 205 participants with non-PsA psoriasis conditions, including psoriasis vulgaris (PsV; weighted mean age: 50.6 years; 6 females; weighted PASI: 17.7), cutaneous psoriasis (PsC; weighted mean age: 40.4 years; 20 females; weighted disease duration: 10.0 years; weighted PASI: 2.8), and unspecified psoriasis without documented inflammatory arthritis (weighted mean age: 42.0 years; weighted disease duration: 12.9 years; weighted PASI: 18.5). Female counts for unspecified psoriasis were considered not reportable as one large (psoriasis n = 111) study [14] did not separately provide sex distributions for individuals with psoriasis and PsA. The weighted disease duration for individuals with PsV was considered not reportable since both studies that recruited PsV participants [13, 16] did not report disease duration. Additionally, 50 participants possessed other inflammatory joint disorders, including 35 with rheumatoid arthritis (RA; weighted mean age: 58.0 years; 80.0% female; weighted DAS: 4.8) and 15 with gouty arthritis (weighted age NA; 6.7% female). Most studies included middle-aged (>40 years old) adults with psoriasis, while only one study included patients aged < 25 and patients aged 26-40 years [13].

**Figure 2.**
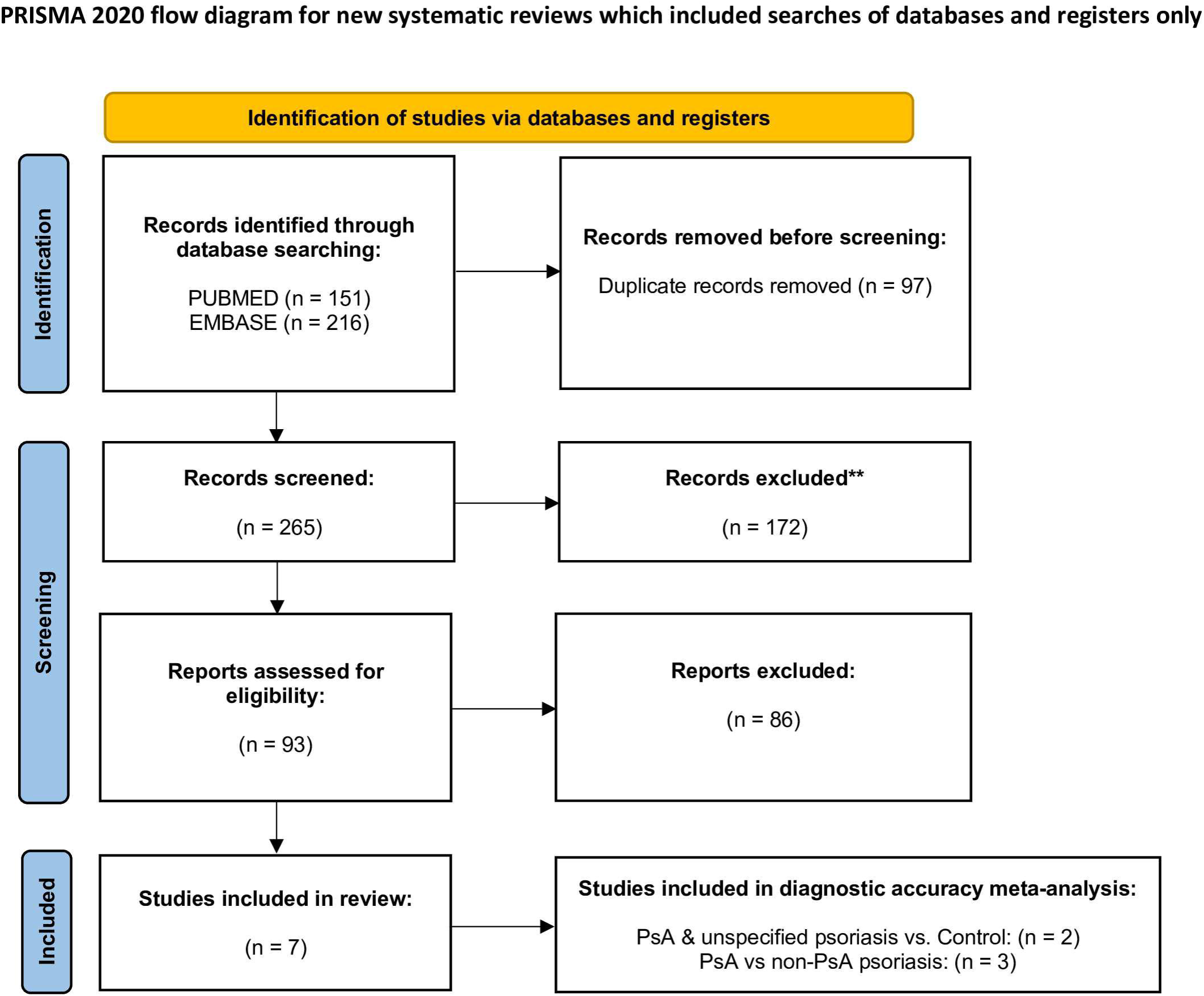
PRISMA flow diagram. EVs were primarily isolated from plasma (n=6 studies) with one study [16] utilizing serum. EV isolation methods varied across studies and included ultracentrifugation (n=2 studies), polymer-based precipitation through ExoQuick and miRCURY exosome isolation kits (n=2 studies) and size exclusion chromatography (n=1 study). Of the two studies using ultracentrifugation, one included a filtration step prior to centrifugation. One study [11] used flow cytometry to characterize circulating EVs using Annexin V in combination with CD31 (for endothelial) and CD41a (for platelets) surface markers. EV characterization techniques included flow cytometry (n=1 study), nanoparticle tracking analysis (n=3 studies), scanning electron microscopy (n=1 study), transmission electron microscopy (n=2 studies), tunable resistive pulse sensing (n=1 study) and western blotting (n=4 studies). Biomarker measurement techniques included flow cytometry (n=3 studies), multiplexed microarray (n=1 study) and ELISA (n=1 study) for proteomic biomarkers. For genetic biomarkers, measurement techniques included miRNA sequencing (n=4 studies) and qPCR (n=1 study). One study [16] also used nanoparticle tracking analysis to evaluate the diagnostic potential of the differences in EV size distribution between disease groups. Of the four studies included in the ROC meta-analysis [11, 15–17], three [11, 15, 16] required data estimation using WebPlotDigitizer. Among these, two studies [11, 16] provided plots with individual-level biomarker data points which were used to approximate biomarker values and one study [15] provided ROC curves which were used to estimate sensitivity and specificity.

### Risk of bias assessment

Studies were classified as cross-sectional or longitudinal, and methodological quality was assessed using modified Newcastle-Ottawa Scales adapted for each study type. Overall, studies demonstrated fair methodological quality, with recurrent limitations related to sample representativeness, EV confirmation, pre-analytical handling, and adjustment for confounding variables. Among the 6 cross-sectional studies (**Table S2**), 3 studies included sufficiently representative samples of individuals with PsA across varying severities and disease activity, while 3 studies selected controls from the same hospital population as exposed participants. Four studies had adequate ascertainment of exposure, with PsA diagnoses established using CASPAR criteria. Across all cross-sectional studies, adjustment for variables such as age, sex, body mass index (BMI), or disease severity was generally unclear or absent. Four studies utilized targeted analytical methods, including qRT-PCR or flow cytometry, to measure EV biomarkers, and 4 studies confirmed EV isolation using ≥2 characterization techniques. Only 2 studies adequately described pre-analytical EV handling procedures, including factors such as fasting status, collection timing, collection tubes, platelet depletion, freezing conditions, or lysis methods. For the single longitudinal study (**Table S3**), selection of controls, demonstration of initial absence of exposure, assessment of EV biomarker outcomes, follow-up duration, and adequacy of follow-up were considered high quality. However, the study lacked a representative PsA sample, confirmation of EV isolation using ≥2 techniques, sufficiently described pre-analytical EV handling procedures, and adjustment for potential confounding variables. Overall, these findings indicate moderate methodological quality, although heterogeneity in EV processing, characterization, and confounder control may affect reproducibility and interpretation of results.

### Diagnostic protein biomarkers

The distinguishing capabilities of general EV surface protein markers among PsA, other psoriasis conditions and controls was heterogeneous (**Table 2**). Two studies [11, 12] utilizing flow cytometry detected significant differences in specific surface marker-positive EVs between individuals with PsA and controls. Ho et al. [11] compared levels of endothelial-cell-derived (Annexin-V+CD31+) and platelet-derived (Annexin-V+CD41a+) EVs in plasma samples of individuals with psoriasis (PsA, n = 3; unspecified psoriasis, n = 8). The authors reported increased levels of endothelial-cell-derived (Annexin-V+CD31+) and platelet-derived (Annexin-V+CD41a+) EVs in psoriasis compared to controls. Meanwhile, Marton et al. [12] compared levels of CD3+, CD14+, CD15+, CD19+, CD42b+, CD235a+, OPG+, RANK+, and RANKL+ EVs within larger-sized EV fractions and smaller-sized EV fractions isolated from plasma samples of individuals with PsA, RA, and controls via differential ultracentrifugation. Within the larger-sized EV fraction, individuals with PsA exhibited increased levels of T cell-derived (Annexin V+CD3+) and granulocyte-derived (Annexin V+CD15+) EVs compared to controls. Within the smaller-sized EV fraction, individuals with PsA exhibited increased levels of erythrocyte-derived (Annexin V+CD235a+) EVs compared to controls, increased monocyte-derived (Annexin V+CD14+) EVs compared to RA, decreased T cell-derived (Annexin V+CD3+) EVs compared to RA, and decreased RANK+ (Annexin V+RANK+) EVs relative to both RA and controls. No significant differences were observed among the remaining markers.

When evaluating EV-associated inflammatory cytokines and protein cargo, Jacquin-Porretaz et al. [14] evaluated EV-associated cytokines (IL-1β, IL-2, IL-6, IL-10, IL-17A, and TNF-α) and heat shock protein 70 (HSP70) in plasma-derived EVs from individuals with mild psoriasis (PsA, n = 3; unspecified psoriasis, n = 46) and moderate-to-severe psoriasis (PsA, n = 6; unspecified psoriasis, n = 65), including participants receiving immunosuppressive therapies such as IL-17 inhibitors (secukinumab, ixekizumab), TNF inhibitors (adalimumab, etanercept), IL-12/23 inhibitors (ustekinumab), conventional disease-modifying anti-rheumatic drugs (methotrexate), and phosphodiesterase-4 inhibitors (apremilast). Following EV isolation via ultracentrifugation, nanoparticle tracking showed no significant differences in EV size distribution or concentration between psoriasis severity groups. However, moderate-to-severe psoriasis exhibited significantly higher EV-associated IL-17A levels compared with mild psoriasis (**Table 2**). This difference remained significant among individuals receiving IL-17 inhibitors, while EV-associated IL-17A levels did not differ between individuals receiving IL-17 inhibitors and other systemic therapies. In contrast, EV-associated IL-1β, IL-2, IL-6, IL-10, TNF-α, and HSP70 did not differ based on disease severity. As baseline EV-associated IL-17A levels prior to treatment initiation were not reported, it remains unclear whether EV-associated IL-17A changes in response to immunosuppressive treatment.

### Diagnostic miRNA biomarkers

Four studies evaluated EV-associated miRNAs as diagnostic biomarkers using sequencing-based approaches, with one study performing qPCR validation [15]. Overlap of differentially expressed miRNAs across studies was limited, suggesting substantial heterogeneity (**Table 3**). Among comparisons between PsA and non-PsA psoriasis, only 4.2% (n=4/95) of miRNAs demonstrated overlapping regulation across studies. Similarly, among comparisons between PsA and controls, only 1.5% (n=1/67) of miRNAs overlapped across studies. Despite this heterogeneity, miR-26a-5p, miR-27a-3p, and miR-27b-3p were consistently upregulated across two independent studies [15, 16], while miR-92a-3p was consistently downregulated [15, 16]. miR-192-5p expression was inconsistent across two studies [13, 17]. Among comparisons with PsA and controls, miR-199a-5p was the only reproducibly upregulated miRNA across two studies [13]. Moreover, qPCR validation identified let-7b-5p and miR-30e-5p as candidate biomarkers as being associated with PsA [15], although these findings lacked replication. Overall, EV-associated miRNA findings demonstrated limited overlap with a small number of reproducible signatures, suggesting that select miRNAs may warrant further investigation as diagnostic biomarkers.

Two studies evaluated the diagnostic or predictive performance EV-associated miRNAs through ROC analysis. Yan et al. [17] assessed miR-218-5p as a biomarker for distinguishing PsA from unspecified psoriasis and reported an AUC of 0.76, with a sensitivity of 73.3% and specificity of 87.5%. Pasquali et al. [15] evaluated let-7b-5p and miR-30e-5p as biomarkers distinguishing PsA from cutaneous psoriasis, reporting AUC values of 0.68 and 0.69, respectively. These miRNAs demonstrated generally low ROC performance (AUC: 0.68-0.76) indicative of limited diagnostic utility. Moreover, Pasquali et al. [15] performed Pearson’s correlations between EV-associated let-7b-5p and miR-30e-5p expression and disease severity measures, including PASI, however no significant correlations were reported (**Table 3**). Additionally, the authors conducted logistic regression adjusted for age, sex, and PASI, reporting that both let-7b-5p and miR-30e-5p predicted lower odds of PsA classification. [15]

### Diagnostic accuracy of EV biomarkers

To identify the best diagnostic biomarker(s) for distinguishing PsA and unspecified psoriasis from controls, as well as PsA from non-PsA psoriasis, we pooled available studies using a BRMA model (**Figure 3A-D**). The model for PsA and unspecified psoriasis vs. controls showed high diagnostic accuracy (AUC = 0.95, sensitivity = 65.0%, specificity = 100.0%). However, due to the small study count (n = 2), these results remain preliminary, and heterogeneity could not be assessed. The model for PsA vs. non-PsA psoriasis showed moderate diagnostic accuracy (AUC = 0.77, sensitivity = 83.0%, specificity = 93.0%) with low heterogeneity (I² = 2.0%), suggesting a promising potential for EV-related biomarkers to distinguish PsA from other psoriasis-related conditions. However, interpretation remains limited by the small number of included studies (n = 3). Notably, one diagnostic miRNA biomarker, miR-26a-5p, which was upregulated in PsA compared to non-PsA psoriasis in two studies [15, 16], also demonstrated strong diagnostic accuracy for distinguishing PsA from PsV (AUC = 0.94, sensitivity = 100.0%, specificity = 87.5%). However, these findings should be interpreted with caution because they were derived from a single study.

**Figure 3.**
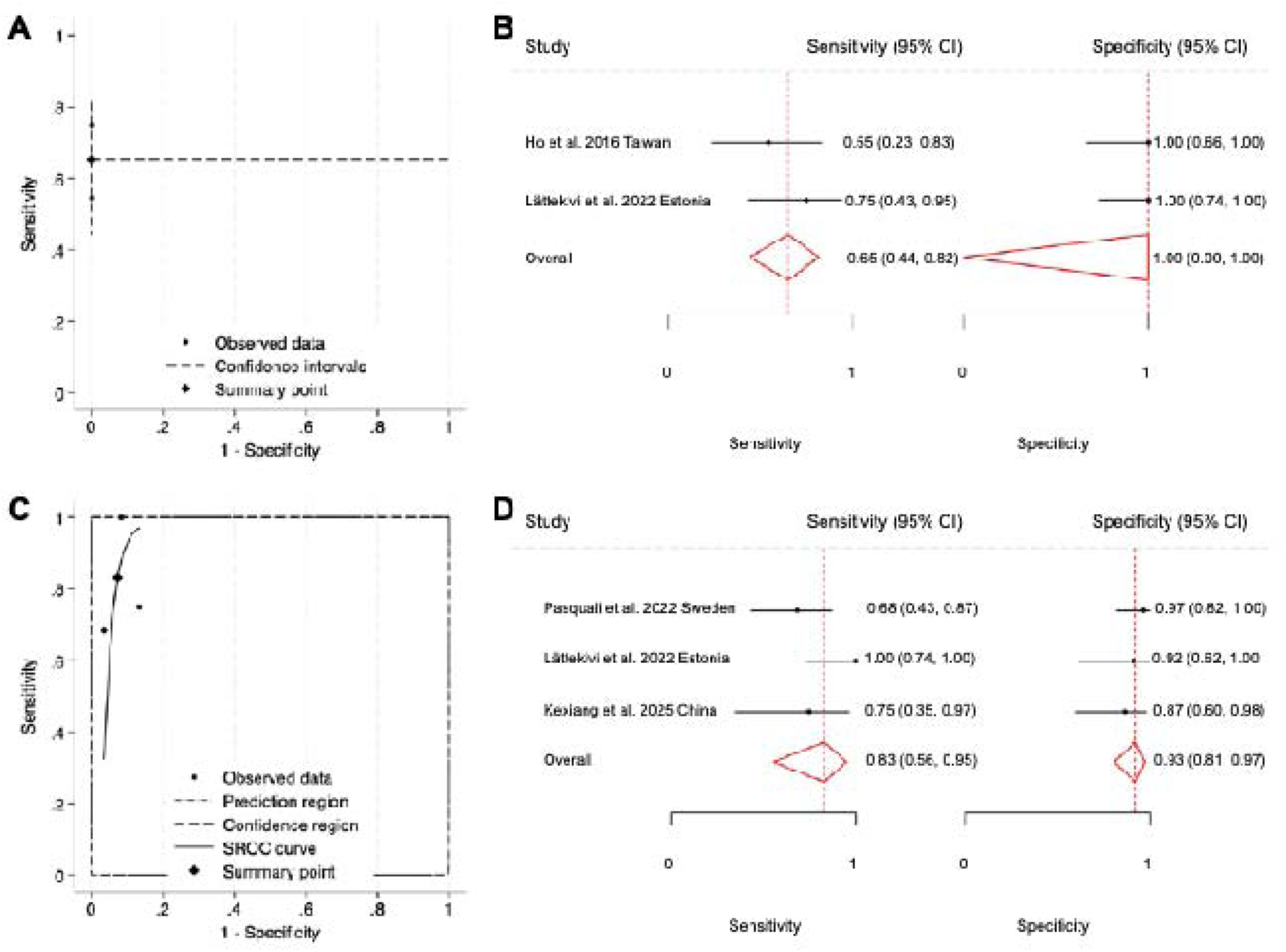
Diagnostic accuracy of extracellular vesicle (EV)-derived biomarkers for distinguishing PsA. (A-B) Diagnostic performance of EV biomarkers for distinguishing PsA and unspecified psoriasis from controls. (A) Summary receiver operating characteristic (SROC) plot showing individual study estimates, confidence intervals, and the pooled summary point. (B) Forest plots of sensitivity and specificity for each included study, with point estimates and 95% confidence intervals shown for sensitivity (left panel) and specificity (right panel); red diamonds and red dashed lines indicate pooled summary estimates. (C-D) Diagnostic performance of EV biomarkers for distinguishing PsA from other psoriasis conditions. (C) SROC curve showing individual study estimates, the summary point, 95% confidence region, and 95% prediction region. (D) Forest plots of sensitivity and specificity for each included study, with pooled summary estimates indicated by red diamonds and dashed lines

### Predictive Biomarkers

Ho et al. [11] evaluated EV biomarkers in a predictive context. The authors analyzed circulating levels of plasma EVs of endothelial (Annexin-V+CD31+) or platelet (Annexin-V+CD41a+) origin obtained from a group of individuals with psoriasis (PsA, n = 3; unspecified psoriasis, n = 8) treated with an anti-IL-12/23 inhibitor (Ustekinumab), at baseline and at a 4-month follow up. No significant differences in Annexin-V+CD31+ or Annexin-V+CD41a+ levels were appreciated following treatment, while the presence or absence of atherosclerosis-related comorbidities did not significantly influence the results.

### EVs as functional biomarkers

Marton et al. [12] examined the role of EVs as potential carriers of functional biomarkers. Ultracentrifugation at 20,500×g and 100,000×g was utilized to isolate larger- and smaller-sized plasma EVs, respectively, from individuals with PsA, RA, and controls. These EVs were then used to treat donor CD14+ monocytes to assess TRAP+ osteoclast formation. As multinucleated TRAP+ osteoclasts are responsible for bone resorption, this model was used to assess whether EVs promote or inhibit osteoclastogenesis, enabling determination of potential contribution to the abnormal bone remodeling and erosive pathology observed in PsA. The smaller-sized EVs derived from healthy controls or RA patients significantly inhibited osteoclast differentiation, while larger-sized EVs did not demonstrate the same inhibitory effect. However, PsA-derived smaller-sized EVs promoted monocyte differentiation to TRAP+ cells. Furthermore, qPCR analysis demonstrated reduced expression of osteoclast-associated genes including *CALCR*, *CTSK*, and *RANK* following smaller-sized EV treatment only in RA and control CD14+ monocytes. These results suggest that smaller-sized PsA-derived EVs may carry biologically active cargo with greater pro-osteoclastogenic or pathogenic potential, promoting osteoclast differentiation. As EVs function as mediators of intercellular communication [24, 25], these findings suggest a potential pathological role for EVs in PsA progression and aberrant bone remodeling through osteoclastogenesis.

## Discussion

Early detection and diagnosis of PsV remains difficult secondary to overlap with other inflammatory arthropathies and the absence of highly specific molecular biomarkers, contributing to delayed diagnosis, leading to progressive radiographic changes and decreased functional outcomes at the time of intervention [1]. Given the emerging role of EVs as minimally invasive biomarkers reflecting inflammatory and immune signaling, this review sought to systematically review the current literature evaluating EV-associated proteins and miRNAs in PsA. Across 7 included studies (**Table 1**), EV-associated proteins demonstrated heterogeneous findings related to inflammatory, vascular, and osteoimmunological pathways (**Table 2**), while EV-associated miRNAs demonstrated substantial heterogeneity with limited overlap across studies. However, a very small number (n=4/95) of reproducible miRNA signatures were identified (**Table 3**). EVs represent a promising set of biomarkers in PsA with potential function as mediators of disease processes through biologically active cargo involved in inflammation and abnormal bone remodeling [12]. However, methodological heterogeneity, limited reproducibility, and insufficient longitudinal evidence warrant further investigation

Together, EV-associated protein and miRNA findings suggest that PsA may be characterized by coordinated changes in inflammatory, vascular, synovial, and skeletal remodeling pathways. Compared with controls, individuals with PsA demonstrated increased levels of EVs derived from endothelial cells (CD31+), platelets (CD41a+), T-cells (CD3+), granulocytes (CD15+), and erythrocytes (CD235a+), potentially reflecting the characteristic heightened systemic inflammation, immune activation, vascular dysfunction, and increased cellular turnover associated with psoriatic disease [11, 12]. Notably, PsA exhibited reduced levels of RANK+ EVs relative to both controls and RA despite elevated EV populations from several inflammatory cell types associated with osteoclastogenesis [12]. This observation aligns with previous findings from Diani et al. [26] who reported reduced serum RANKL concentrations in individuals with PsA compared with controls, suggesting that alterations in RANK/RANKL-related signaling may extend across both circulating proteins and EV-associated markers. Increased miR-199a-5p in PsA compared with controls may provide further implications on cartilage and skeletal remodeling pathways [13, 17], as prior evidence has shown that miR-199a-5p may promote osteoclast differentiation through mechanisms distinct from the RANK/RANKL pathway [27].

In EV treatment experiments, PsA-derived EVs did not significantly alter the expression of osteoclast-related markers, including *CALCR*, *CTSK*, and *RANK*, in CD14+ cells, whereas RA-and control-derived EVs showed inhibitory effects, suggesting that PsA-derived EVs may lack the same regulatory influence on RANK/RANKL-associated osteoclastogenic signaling observed with RA- and control-derived EVs. Additionally, increased EV-associated IL-17A levels in individuals with more severe psoriasis may support the importance of the IL-23/IL-17 inflammatory axis in psoriatic disease progression [14]. The investigation by Suzuki et al. [28] reported that IL-23/IL-17 signaling may promote RANK-associated inflammatory pathways, while also potentially driving osteoclastogenic mechanisms independently of RANK/RANKL signaling. Together, these findings suggest that although PsA may be characterized by enhanced inflammatory and immune cellular activity, osteoclastogenic and bone remodeling processes may not depend primarily on upregulation of the RANK/RANKL pathway.

EV profiles may also help distinguish PsA from clinically overlapping inflammatory diseases, including RA and non-PsA psoriasis. In comparisons with RA, PsA was characterized by increased monocyte-derived (CD14+) EVs and reduced T-cell-derived (CD3+) EVs, suggesting that EV cellular origin may reflect immune differences between these two arthritic conditions [12]. Separately, EV-associated miRNA comparisons between PsA and non-PsA psoriasis suggest that miRNA cargo may help distinguish arthritic from cutaneous-specific psoriatic disease. In these comparisons, miR-26a-5p, miR-27a-3p, and miR-27b-3p were consistently upregulated in PsA and linked to fibroblast-like synoviocyte activation, inflammatory signaling, apoptosis regulation, and invasive synovial behavior in inflammatory arthritis models [15, 16]. These processes are directly relevant to joint inflammation and structural damage, suggesting that these miRNAs may reflect arthritic rather than purely cutaneous disease activity. In contrast, reduced miR-92a-3p in PsA compared with non-PsA psoriasis may indicate a shift away from skin-dominant keratinocyte inflammatory programs, as the miR-17–92 cluster has been associated with keratinocyte proliferation and inflammatory signaling in psoriasis [15, 16].

Together, these findings suggest that EV-associated cellular and miRNA signatures may help differentiate PsA from both RA and non-PsA psoriasis by possessing distinct patterns of immune activation, synovial involvement, and cutaneous inflammation. Consistent with this, our diagnostic accuracy meta-analysis for PsA vs. non-PsA psoriasis showed moderate pooled performance, supporting the potential diagnostic relevance of EV-related biomarkers in distinguishing arthritic from non-arthritic psoriatic disease. Among these candidates, miR-26a-5p appears particularly promising, as it was upregulated in PsA compared to non-PsA psoriasis across two studies [15, 16] and showed a strong estimated diagnostic signal for distinguishing PsA from PsV in one study [16]. However, these findings remain preliminary because the diagnostic evidence is based on a small number of studies and includes digitized data estimations. Future studies should combine EV protein and miRNA profiles into multi-modal biomarkers, which may improve differentiation of PsA from RA and non-PsA psoriasis by capturing complementary inflammatory and immune signatures, while also enabling stronger validation of the preliminary diagnostic signals observed in ROC-based analyses.

This review is not without limitations. The number of included studies was small, and study populations were heterogeneous with respect to disease severity, psoriasis subtype, treatment status, and comparator groups, limiting the generalizability of the findings and making PsA-specific interpretation difficult. In some studies, PsA was grouped within broader psoriasis cohorts, while only four studies used CASPAR criteria to define PsA [12, 13, 15, 17], raising the possibility that some findings may not fully reflect true PsA-specific pathology. In addition, based on reported PASI scores, many studies did not capture a broad range of disease severity, further limiting conclusions about how EV-associated markers may vary across clinical phenotypes. Methodological variability in EV isolation, characterization, and molecular profiling also likely contributed to inconsistent findings across studies, including differences in plasma versus serum sampling, ultracentrifugation, polymer-based precipitation, size exclusion chromatography, and sequencing approaches. Furthermore, a number of studies did not adjust for potential confounders such as age, sex, BMI, treatment exposure, and disease severity. These limitations are reflected in the biomarker results, where limited overlap across studies, modest ROC performance of markers such as let-7b-5p and miR-30e-5p [15], and inconsistent regulation of miRNAs such as miR-192-5p were appreciated, [13, 17] suggesting that EV-associated signatures remain heterogeneous and sensitive to study design, sample source, comparator group, and disease phenotype. The diagnostic accuracy meta-analyses should also be interpreted cautiously because both ROC models were based on very small numbers of studies, limiting the precision and stability of pooled sensitivity, specificity, and AUC estimates, as well as the ability to robustly assess between-study heterogeneity. In addition, several diagnostic inputs were not directly reported by the original studies but were manually estimated from published figures using WebPlotDigitizer. Although this approach enabled inclusion of otherwise unavailable data, hand-digitized estimates may be affected by plot resolution, axis scaling, point overlap, visual extraction error, and reconstruction of individual-level values or ROC coordinates from non-original data. Finally, as most investigations were cross-sectional in design, it remains unclear whether EV biomarkers are predictive of PsA onset, progression, or treatment response. Therefore, while current findings support a potential role for EV-associated markers as biologically informative of PsA-related synovial and skeletal pathology, EVs should not be relied upon as a standalone diagnostic signature. Future studies should prioritize larger, well-characterized longitudinal cohorts with standardized EV methodologies, CASPAR-defined PsA populations, and more homogeneous comparator groups to better validate candidate biomarkers in determining the diagnostic and predictive utility of EVs.

## Conclusion

EV-associated proteins and miRNAs are potential biomarker candidates for PsA, reflecting inflammatory, vascular, and osteoimmunological processes underlying disease pathophysiology. However, current evidence remains preliminary and limited by small cohorts, methodological heterogeneity, and inconsistent reporting across studies. Larger standardized longitudinal studies are warranted to validate candidate EV biomarkers, determine their predictive value in individuals with psoriasis at risk for PsA, while clarifying whether EV-based approaches improve early diagnosis, disease monitoring, and future therapeutic development.

## Data Availability

Data analyzed are included in the respective figures.

## Authorship contributions

HBT conceptualized and designed the study. HBT & TZ wrote the manuscript. TZ, FSZ, CZ, JA, JL, SN, ES & HBT collected data and checked it for accuracy. SW, RHB & DMK edited and provided critical comments on the manuscript. All authors approved the manuscript submission.

## Acknowledgement

None

## Funding

None

## Conflict of interest

R.H.B.: support for education and hospitality payments from Elite Orthopaedics and hospitality payments from Zimmer Biomet D.M.K.: support for education from Synthes, Smith & Nephew, Elite Orthopedics, and Medwest Associates; hospitality payments from Arthrex, Elite Orthopaedics, Encore Medical, Stryker, and Smith & Nephew; honoraria from Encore Medical; and a grant from Arthrex. Other authors report no conflict of interest.

## Notes

### Author Declarations

This is a systematic review & meta-analysis of publicly available data from the respective studies.

