## Supplementary material for "Extracellular vesicles as biomarkers for psoriatic arthritis: a systematic review & meta-analysis": Table S1

**Table S1: Complete Search strategy using PUBMED and EMBASE. The search was performed from the date of inception until May 21^st^, 2026.**

| PUBMED | ( "extracellular vesicle*"[Title/Abstract] OR exosome*[Title/Abstract] OR ectosome*[Title/Abstract] OR microvesicle*[Title/Abstract] OR "apoptotic bod*"[Title/Abstract] ) AND ( psoriasis[Title/Abstract] OR psoriatic[Title/Abstract] OR "psoriatic arthritis"[Title/Abstract] OR "plaque psoriasis"[Title/Abstract] OR "guttate psoriasis"[Title/Abstract] OR "skin inflammation"[Title/Abstract] OR "keratinocyte proliferation"[Title/Abstract] ) NOT ( review[Publication Type] OR systematic review[Publication Type] OR meta-analysis[Publication Type] ) |
| --- | --- |
| EMBASE | ( 'extracellular vesicle'/exp OR exosome/exp OR microvesicle/exp OR 'extracellular vesicle*':ti,ab OR exosome*:ti,ab OR microvesicle*:ti,ab ) AND ( psoriasis/exp OR 'psoriatic arthritis'/exp OR psoriasis:ti,ab OR psoriatic:ti,ab OR 'plaque psoriasis':ti,ab OR 'guttate psoriasis':ti,ab ) NOT ( review/it OR 'systematic review'/it OR 'meta analysis'/it ) |
